# A downscaling approach to compare COVID-19 count data from databases aggregated at different spatial scales

**DOI:** 10.1101/2020.06.17.20133959

**Authors:** Andre Python, Andreas Bender, Marta Blangiardo, Janine B. Illian, Ying Lin, Baoli Liu, Tim Lucas, Siwei Tan, Yingying Wen, Davit Svanidze, Jianwei Yin

**Affiliations:** Center for Data Science, Zhejiang University, 866 Yuhangtang Road, Hangzhou 310058, Zhejiang Province, P. R. China; Big Data Institute, Li Ka Shing Centre for Health Information and Discovery, University of Oxford, Old Road Campus, Oxford, United Kingdom; School of Mathematics and Statistics, Mathematics and Statistics Building, University of Glasgow, University Place, Glasgow G12 8QQ, United Kingdom; Department of Statistics, LMU Munich, Akademiestrasse 1/IV, 80799 Munich, Germany; Department of Epidemiology and Biostatistics, St Mary’s Campus, Imperial College London, United Kingdom; College of Environment and Resources, Fuzhou University, 2 North Wulong Street, 350108 Fuzhou, P. R. China; School of Geography and the Environment,University of Oxford, South Parks Road, Oxford, OX1 3QY, United Kingdom; Department of Statistics and Econometrics, Georg-August-Universität Göttingen, Wilhelmsplatz 1, 37073 Göttingen, Germany

**Keywords:** COVID-19, downscaling, spatially disaggregated data

## Abstract

As the COVID-19 pandemic continues to threaten various regions around the world, obtaining accurate and reliable COVID-19 data is crucial for governments and local communities aiming at rigorously assessing the extent and magnitude of the virus spread and deploying efficient interventions. Using data reported between January and February 2020 in China, we compared counts of COVID-19 from near-real time spatially disaggregated data (city-level) with fine-spatial scale predictions from a Bayesian downscaling regression model applied to a reference province-level dataset. The results highlight discrepancies in the counts of coronavirus-infected cases at district level and identify districts that may require further investigation.

## 1. Introduction

By the end of December 2019, a pneumonia of unknown cause was identified in Wuhan, Hubei province. The World Health Organization (WHO) temporarily named the cause of the pneumonia severe acute respiratory syndrome coronavirus 2 (SARSCoV2). The disease associated with the syndrome took the name coronavirus disease 2019 or COVID19 ^[1–3]^. On the 30^th^ of January 2020, the WHO declared the outbreak of COVID19 a Public Health Emergency of International Concern, the highest level of international emergency response given to infectious diseases^[4]^. By June 9^th^ 2020, COVID-19, had spread worldwide, infected more than 7 million individuals, and had led to about 400,000 deaths (confirmed cases that died) worldwide^[5]^.

Efforts have been made by various institutions to collect near-real time data at city level (disaggregated data) on COVID-19 worldwide and to make it publicly available^[6–9]^. While there is a race against time to contain the outbreak, reliability and accuracy of COVID-19 data should remain a top priority for the research community. Policymakers require evidence-based analysis to design and implement efficient interventions to mitigate and prevent the spread of the virus. A key condition for efficient evidence-based interventions is to ensure that policy recommendations are based on reliable data measured with sufficient accuracy^[10]^.

COVID-19 is part of a large family of coronaviruses, which seems to have transferred from animals to humans between November and December 2019, according to the phylogeny of genomic sequences obtained from early cases^[11]^. Recent work showed that COVID-19 can transfer quickly and easily among individuals^[12]^. While the symptoms are often mild, the virus seems to be more lethal for old people and individuals with preexisting medical conditions^[4]^. A large number of countries are highly vulnerable to a COVID-19 epidemic. Globally, only 50% of countries have a national infection prevention and control program and about 30% of countries have no COVID-19 national preparedness and response plans^[13]^.

The establishment of containment areas, production and distribution of respiratory masks, and the delivery of urgent medical care tend to be very costly^[4,14]^. To improve the efficiency of interventions, and hence, reduce the number of infected individuals and associated fatalities, epidemiological models based on geolocalized data can be employed to: (a) make inference on the true number of cases in spatial locations; (b) investigate the mechanisms behind the spread of the virus; (c) identify locations at risk; (d) quantify and forecast the spread of the virus at fine spatial scales to help decision makers target interventions.

Epidemiological models using geolocalized data require sufficient accuracy with regard to both the number counts and the spatial locations of the reported cases. In order to assess data accuracy, we compare two major providers of spatially disaggregated COVID-19 data with province-level data from Johns Hopkins University Center for Systems Science and Engineering (JHU)^[15]^. An exploratory analysis based on reported data in China from January to February 2020 indicates that both the number of corona-infected cases from disaggregated datasets and the spatial locations of the reported cases exhibit important divergences between the investigated datasets. We further investigate differences in the counts at a fine spatial scale (district-level) between reported values from spatially disaggregated datasets and values estimated from a Bayesian downscaling modeling approach.

The downscaling approach presented in this paper estimates the counts of COVID-19 and its uncertainty at fine-scale (5 km grid-cells) using spatially aggregated (province-level) data and covariates (5 km grid-cells) as input. Predictions from the downscaling model are compared at district-level with COVID-19 from spatially disaggregated datasets. The results of this analysis suggest that COVID-19 counts reported by the disaggregated datasets are often consistent with the range of plausible estimated counts from the downscaling model, however data coverage and discrepancies in the counts remain important in various districts. The resulting maps can be visualized in an ESRI dashboard (https://arcg.is/008GDX).

## 2. Method

### 2.1. Data

As an initial exploratory analysis, we examine two fundamental metrics on COVID-19: the number of confirmed COVID-19 infected cases and the locations of the reported cases. We investigate two major providers of COVID-19 spatially disaggregated data which provide the location (longitude, latitude) of cities and the date of reported coronavirus-infected cases. We compare two disaggregated datasets: (1) *Xu et al*. data^[16]^ and (2) *The Paper* data. *Xu et al*. data has been published in *Scientific Data* ^[16]^ and publicly available through GitHub repository^[8,9]^. The Paper data is provided by the Pengpai news agency^[6]^, also refers to as *The Paper*. The disaggregated data from the two providers are regularly updated online. In this case study, we compare them with province-level data provided by Johns Hopkins University Center for Systems Science and Engineering (JHU)^[15]^.

First, we compared the count of confirmed COVID-19 infected cases reported from January to February 2020 in China at province level. For consistency, we used data that has been updated by the investigated databases on the same day (April 9, 2020). In Hubei province—the Chinese province most affected by coronavirus—the reported number of COVID-19 infected cases shows large differences among the databases: JHU (65,914 cases), The Paper (58,292 cases), and Xu et al. (20,336 cases).

For the sake of readability, we split the results into two groups: *high-impacted* provinces counting a maximum number of COVID-19 cases (all providers combined) larger than the median (Fig 1, *left panel*) and *low-impacted* provinces counting a maximum number of COVID-19 cases (all providers combined) smaller or equal to the median (Fig 1, *right panel*). We observe important discrepancies in other provinces (Fig 1) between the reported cases provided by The Paper and Xu et al. with JHU data. This may suggest that the providers use different collection methodologies and control processes to gather and select the cases.

**Figure 1:**
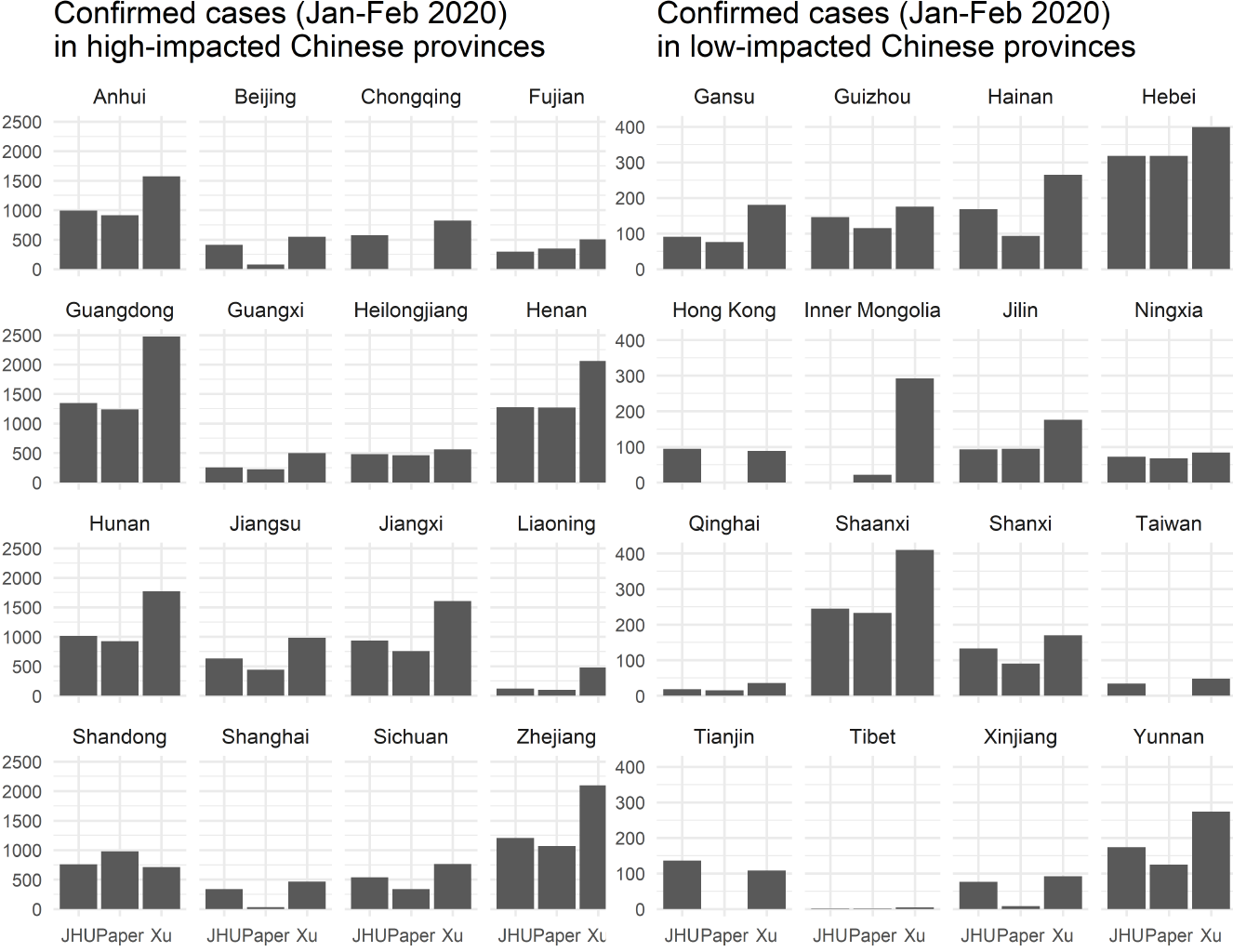
Cases of coronavirus in high-impacted (*left panel*) and low-impacted (*right panel*) provinces in China. The plot indicates the number of coronavirus-infected individuals in China in high-impacted (*left panel*) and low-impacted (*right panel*) provinces from January to February 2020. Note that data from Macau is included in Guangdong province. Sources (updated April 9, 2020): (*left bar*) Johns Hopkins University CSSE^[7]^, (*center bar*) The Paper^[6]^, and (*right bar*) Xu et al. ^[16]^.

Second, we examine spatial variability of the data measured as the number of unique spatial coordinates associated with the reported COVID-19 cases. JHU is excluded from this comparison since it does not provide spatial coordinates along with the reported cases. Therefore, we compared the number of unique locations and associated observed cases provided by Xu et al. and The Paper only. We illustrate the results for cases reported in Hubei province. In this province, The Paper (Fig 2, *left panel*) provides nine locations with reported cases. Fig 2 (*right panel*) shows that the number of locations that reported cases is systematically higher in Xu et al. compared to the Paper data (except in Hebei province).

**Figure 2:**
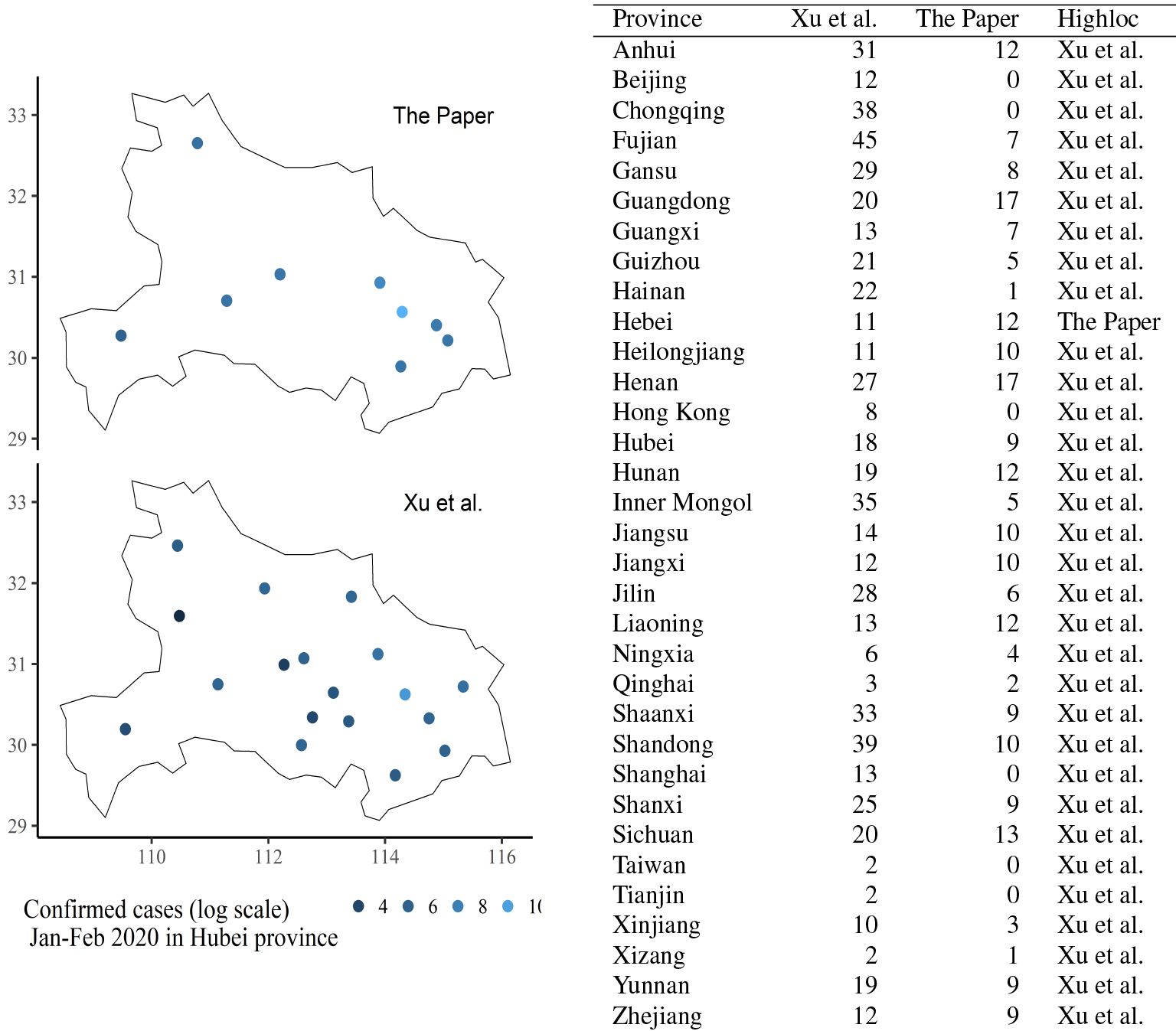
Spatial accuracy of disaggregated datasets. *Left panel*: geolocalization and associated counts (in natural logarithm scale) of coronavirus-infected individuals in Hubei from January to February 2020. *Right panel*: number of locations corresponding to COVID-19 cases (January and February 2020) reported by Xu et al. and The Paper in Chinese provinces. The column *Highloc* indicates the data source that has the largest number of locations with data. Sources (updated April 9, 2020): `The Paper`^[6]^ (*left panel*), and `Xu et al`. (*right panel*)^[16]^.

### 2.2. Downscaling COVID-19 province-level data

To further investigate the observed discrepancies among COVID-19 data, we use a downscaling approach to predict at a fine spatial scale the expected cases from data provided by JHU^[15]^, a major reference for national and subnational data on COVID-19, which provides data consistent with daily data from the Chinese centre for disease control and prevention and WHO situation reports^[15]^. We use the R package `disaggregation` which implements in R a Bayesian approach to carry out spatial disaggregation modeling, also called downscaling^[17]^. Similar approaches have been recently used to estimate fine-scale (grid-cell) risk of disease based on spatially aggregated (polygon-level) data^[18–23]^.

We apply a downscaling method to estimate cases of COVID-19 and their uncertainty within fine grid-cells (about 5km spatial resolution) in China from JHU data aggregated at province-level^[15]^. The model is implemented in template model builder (`TMB`)^[24]^), an R package that provides a suitable framework based on `C++` to build and efficiently fit hierarchical Bayesian models, including downscaling models. The model predictions are compared with observed values from the two disaggregated data: Xu et al.^[16]^ and the Paper^[6]^.

Making predictions at a spatial scale finer than the input data is subject to potential validity threats that result from a mismatch between the scale from which the data and the prediction are considered^[25]^. One issue, known as the “ecological fallacy” has been well documented in geography and spatial epidemiology since the early 1950s^[26]^. A classical example is the study of Durkheim (Le Suicide, 1897), whose analysis showed that suicide rates in Prussia were highest in provinces that were heavily Protestants and wrongly concluded that stronger social control among Catholics would result in lower suicide rates. The author did not envision that it may have been non-Protestants (primarily Catholics) who were committing suicide in predominantly Protestant provinces^[27]^. Analogously, an observed relationship between average population density and the risk of COVID-19 at province-level may not hold at city-level.

A second issue is associated with the effects of the choice of spatial units on the results. This potential validity threat refers to the “modifiable areal unit problem” (MAUP), well known by geographers and epidemiologists^[28]^. Within a lattice framework which uses polygons defined as the spatial unit, one needs to choose the shape (regular or irregular polygons) and size of the polygons used to make predictions at a desired spatial scale^[29]^. To avoid the risk of ecological fallacy and mitigate the effect of the MAUP, the downscaling approach used here takes benefit of the spatial information gathered from fine-scale covariates to model the underlying processes behind the observed data that explain the spatial variations of COVID-19 counts within provinces.

We model the count of coronavirus-infected cases *y*_*i, j*_ for each 5 km grid-cell *j* that lies within each Chinese province (polygon *i*), with *N* the total number of provinces. A continuous-space data-generating process is discretized into 5 km grid-cells from which data is aggregated and estimates are generated. In this framework, we consider the total count of COVID-19 in a given province *y*_*i*_ as the sum of each individual count *y*_*i, j*_ in all grid-cells that cover province *i*. We model the incidence rate, a relevant risk metric of COVID-19 that is defined as the number of new coronavirus-infected during our investigated period (January-February 2020), divided by the population at the beginning of the period.

The incidence rate *λ*_*i, j*_ in pixel *j* of polygon *i* is associated via a log link function to a linear predictor *η*_*i, j*_ composed of an intercept *β*_0_, *N*_*q*_ covariates 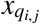 with coefficients *β*_*q*_, a spatial random field *ζ*(*i, j*), and an i.i.d random effect *u*_*i*_ associated with each province (*i* = 1, …, *N*):

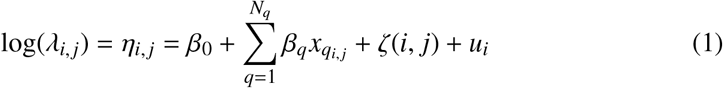

At polygon-level, the conditional distribution of the number of coronavirus-infected individuals approximately follows a Poisson distribution *y*_*i*_ ∼ *Pois* (µ_*i*_), with expected cases define as 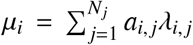, with weight *a*_*i, j*_ used to estimate COVID-19 counts in each polygon. The predicted sum of the counts from each grid-cell over a province is weighted by the population size values in the corresponding grid-cell extracted from a population raster (see Fig 3)^[30]^. The estimated incidence rate corresponds to the expected cases divided by the weights, with 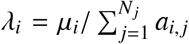. Moreover, *λ*_*i, j*_ is linked to the linear predictor through a log function so that one can derive the incidence rate as *λ*_*i, j*_ = exp(*η*_*i,j*_), where *η*_*i, j*_ is the linear predictor, observed in a bounded spatial window D delimited by the Chinese national border.

**Figure 3:**
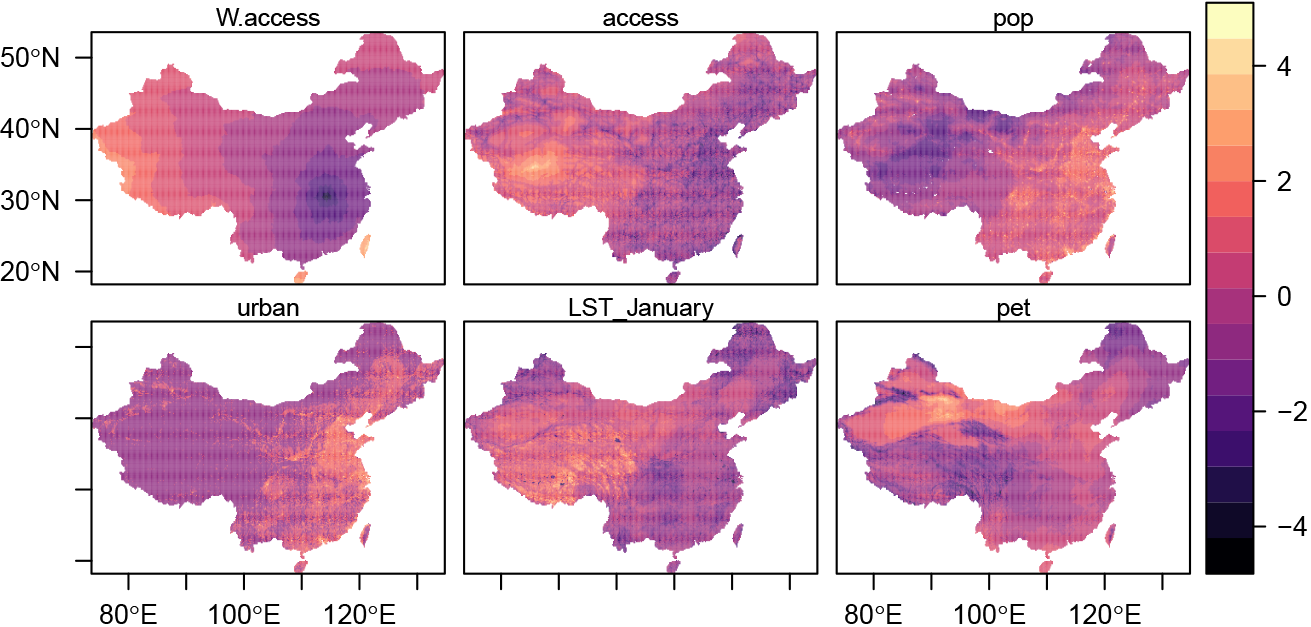
Potential covariates used in the downscaling model. The maps show the standardized values of six potential covariates used in the downscaling model. The model includes travel time to Wuhan, China (*W*.*access*), travel time to to the nearest city with more than 50,000 inhabitants (*access*), population size (*pop*), urbanity (*urban*), January land surface temperatures (*LST*), potential evapotranspiration (*pet*).

The linear predictor includes *N*_*q*_ covariates **x**_*q*_(*i, j*), with coefficients *β*_*q*_ (*q* = 1, …, *N*_*q*_). Covariates associated with COVID-19 infections are numerous and operate at various spatial levels, from the host genetics to large-scale transnational factors that affect travel between countries for example. To make predictions at pixel-level (about 5 km resolution), we gather covariate data from various sources, mainly raster data (covariates aggregated into regular grids) which cover China. Climate and meteorological factors can affect the life cycle of respiratory diseases and their propensity to spread^[31]^. However the extent of the role of climate drivers on the spread of COVID-19 compared to those associated with human behaviour is not well understood^[32]^. COVID-19’s transmission can potentially occur in any ecological context by human contact. Therefore, the main drivers of the transmission of COVID-19 are likely to be associated with human behaviour rather than environmental factors^[33]^. We consider mainly covariate data that are associated with human activity and behaviour and include only a few relevant environmental variables. We also note that we are not making predictions outside of the area or time of study but restrict our analysis to the downscaling of cases in China which avoids many of the main issues associated with using fine-scale covariates.

COVID-19 appears to transfer rapidly and easily among humans, with a basic reproductive number *R*_0_ (also called transmission rate) seemingly between 2 and 3. *R*_0_ represents the expected number of direct infections from one case in a completely susceptible population^[34]^. This means that, one person infected by COVID-19, in a completely susceptible population, transfers on average the virus to 2 or 3 people^[4]^. The basic reproductive rate depends on various factors associated with the virus itself and the risk of individuals transferring the virus, directly or indirectly, to other individuals as well.

We consider six covariates (1-6) with full coverage in China at 5 km resolution (Fig 3). The selected set of covariates include proxies for human activity, population density, and transportation: (1) travel time to the nearest city of with more than 50,000 inhabitants (*access*)^[35]^, (2) population size (*pop*) from WorldPop^[30]^, and (3) urbanity (*urban*) from Global Urban Footprint data^[36]^. Along with population size and urbanity, travel time to the nearest large city is likely to be an important contributor of the spread of coronavirus at pixel-level, since it measures proximity to large urban centers. Locations close to city centers affected by COVID-19 are more likely to be affected by a spread of the disease. Since COVID-19 was first identified in Wuhan, we include (4) travel time to Wuhan, China (*W*.*access*) (computed from^[35]^) to account for a potential diffusion from Wuhan.

We consider two environmental covariates that are likely to have a role in the spread of COVID-19. We include (5): January land surface temperatures (*LST_January*) from the US National Oceanic and Atmospheric Administration for meteorological (NOAA) Geostationary Operational Environmental Satellites (GOES)^[37]^). Temperatures in January can affect the life cycle of COVID-19 and their propensity to spread^[31]^, but can also affect human behaviour. The effects of cold temperatures on human transmission of COVID-19 are not clear. On the one hand, cold weather can increase the risk of transmission as people spend more time indoors, but could also reduce the risk of transmission due to a decrease of contact resulting from a reduction human activity.

There is evidence that humidity affects influenza-type virus transmission in both experimental and observational studies^[38,39]^, and in population-level studies^[40]^. High levels of humidity may provide a suitable framework for an outbreak of COVID-19 and modulate the magnitude of the pandemic^[32]^. To account for the role of humidity, we include: (6) potential evapotranspiration (*pet*), which provides a measure of the potential amount of evaporation (if a water body is present), which is associated with the quantity and lifetime of droplets in the air that can directly affect the spread of the disease in the air, or indirectly (e.g. reduce the protective ability of masks)^[41]^.

To mitigate a potential risk of multicolinearity, we applied a variance-inflation factor function to identify and remove collinear covariates that show high levels of correlation using a stepwise procedure from the R package usdm^[42]^. The procedure removes one covariate from all pairs of covariates that exhibit Pearson’s *ρ* > 0.75). We found that travel time to the nearest city of more than 50,000 inhabitants (*access*)^[35]^ and travel time to Wuhan (Hubei province) are highly correlated (*ρ* > 0.75). We kept travel time to Wuhan since we believe that proximity with Wuhan is a major driver of the spread of COVID-19, especially over the study period (January-February 2020), which corresponds to the early pandemic which has been first identified in Wuhan.

In equation 1, the spatial structure *ζ*(*i, j*) is represented by a zero-mean Gaussian Markov random field (GMRF), while 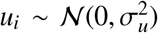 is a polygon i.i.d. random effect, which account for variation among polygons, with variance 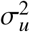. The GMRF has a Matérn covariance function defined by two parameters: the range *ρ*, which represents the distance beyond which correlation becomes negligible (about 0.1), and *σ* is the marginal standard deviation.

Due to the Bayesian setting of the downscaling model, priors need to be defined for all parameters (and hyperparameters) in the model. We used default priors (Normally distributed with mean and standard deviation) for the intercept *β*_0_ ∼ 𝒩(0, 2) and covariate coefficients *β*_*q*_ ∼ 𝒩(0, 0.4). We used penalized complexity (PC) priors^[43]^ for the polygon i.i.d effects *u*_*i*_ ∼ 𝒩(0, τ_*u*_), with standard deviation *σ*_*u*_ (precision 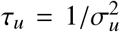). Here, we used the default configuration that favors a base model without polygon-specific effect, with *P*(*σ*_*u*_ > *σ*_*u,max*_) = *σ*_*u,prob*_, with *σ*_*u,max*_ and *σ*_*u,prob*_ that can be defined by the user^[44]^.

For the parameters of the random field (range and scale), we used penalized complexity (PC) priors^[43]^ so that *P*(*ρ* < 3) = 0.01 and *P*(*σ* > 1) = 0.01. Throughout this approach, we constrained the model to favor a random field with a smaller magnitude and a large range. We compare the predictive out-of-sample and in-sample performance of different model specifications, which include variations in the spatial parameters (see details in **SI**).

## 3. Results

We carried out an out-of-sample procedure to compare the predictive performance (MAE, RMSE) of different model specificities by varying several components (see details in **SI**). The investigated models include: (1) all covariates; (2) only anthropogenic covariates; (3) only environmental covariates. In addition, we built four alternative models (4-7) that include all covariates but use alternative penalized complexity (PC) priors on the spatial parameters. The results show that the model including only anthropogenic covariates has in general the highest predictive performance. This is expected, since the spread of COVID-19 is likely to be mainly driven by anthropogenic factors^[33]^.

### 3.1. Parameter estimation

The estimated mean and 95% credible intervals of the fixed effects (Fig 4, *top panel*) are provided for (from bottom to top): the log precision of the i.i.d effects (polygon-level), intercept, spatial hyperparameters log *ρ* and log *σ*, and the *β* coefficients associated with the covariates.

**Figure 4:**
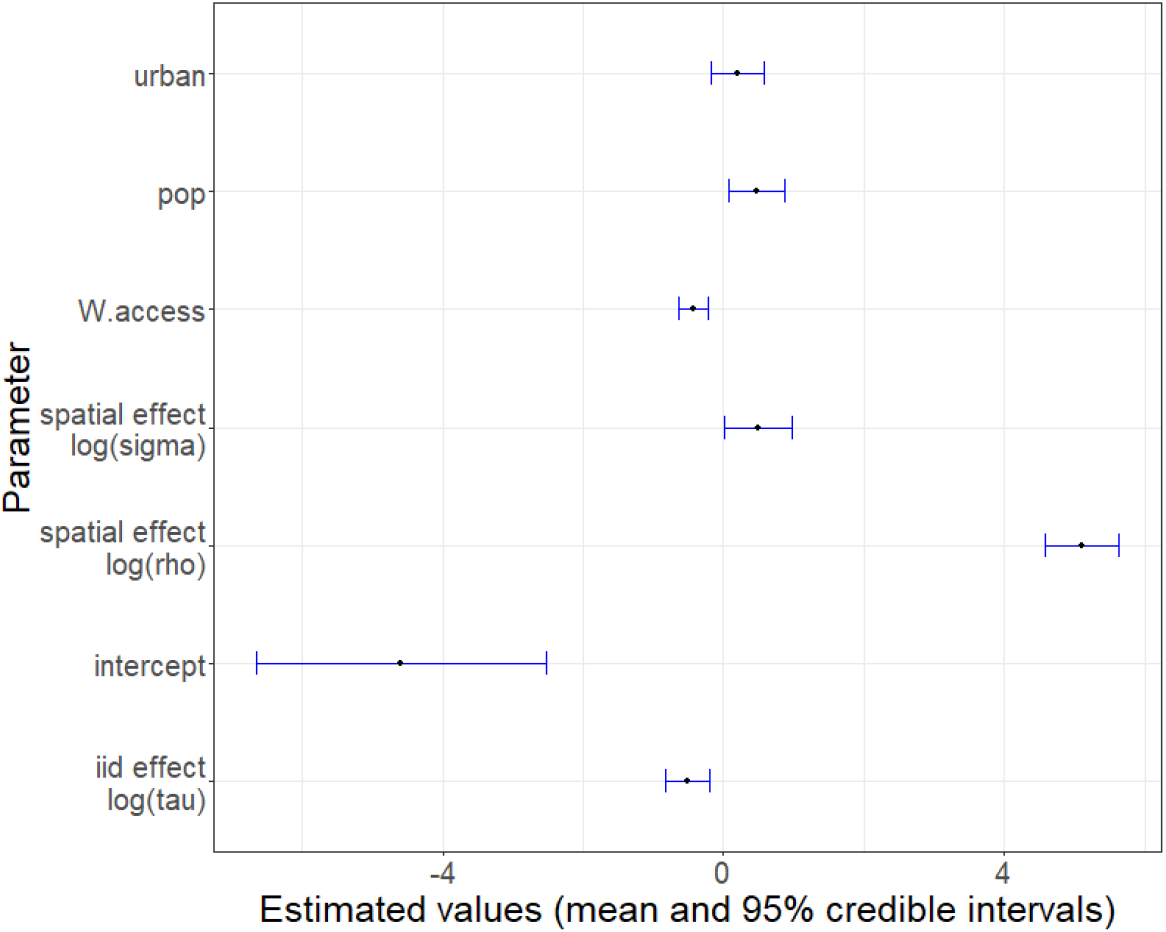
Parameter estimation of the downscaling model. The graphic shows the mean and 95% credible intervals (y-axis) of the estimated parameters (fixed-effects and random effects) in the model (y-axis). It includes (from bottom to top), the log precision of the i.i.d effects (polygon-level), intercept, spatial hyperparameters log *ρ* and log *σ*, and the *β* coefficients associated with each covariate.

As expected, travel time to Wuhan (95% CI: −0.63; −0.22) is negatively associated with COVID-19 incidence rate: the expected COVID-19 incidence rate increases with proximity to Wuhan, where the virus was first identified. We also observed a positive effect of population size (*pop*) on COVID-19 incidence rate (95% CI: 0.08; 0.88), which suggests that areas with larger population size tends to be more prone to COVID-19 infections.

There is evidence of an effect from the i.i.d. province-specific random effects (*log*(τ) (95% CI: −0.82; −0.21), which is expected especially in the early stages of the epidemic, where Hubei province had a much higher number of reported cases compared to the other provinces, whose connection with Wuhan are highly variable can can therefore lead to important risk variability among provinces. A large range of plausible values associated with urbanity are positive, however, the 95% credible intervals of the estimated corresponding coefficient also include a range of negative values (95% CI: −0.18; 0.57). Therefore our results are inconclusive with regard to the role of urbanity on COVID-19 incidence rate. Future studies based on larger datasets might provide additional information on its potential effect.

### 3.2. Downscaling model predictions of COVID-19 cases at fine spatial scales

A sampling process (n=2,000) from the posterior distribution allows us to estimate the expected incidence rate of COVID-19 for January-February 2020 at 5 km grid-cell across China. In addition, we compute the higher and lower bound of the 95% credible intervals estimates of the incidence rate, which provide a measure of uncertainty of the predictions. To compare the results of the downscaling model with the spatially disaggregated datasets (Xu et al. and the Paper), we convert the predicted rates (mean) and their uncertainty (lower bound, and higher bound estimations) into predicted cases by multiplying them with population size for each grid-cell in China.

Fig 5 shows the (natural log) COVID-19 infected cases for January and February 2020 from the original JHU dataset at province-level in China (*top-left*). In addition, it shows an estimation of the (natural log) infected cases based on the downscaling approach with lower bound of the 95% CI (*top-right*), mean (*bottom-left*), and higher bound of the 95% CI (*bottom-right*) predictions at 5 km grid-cell.

**Figure 5:**
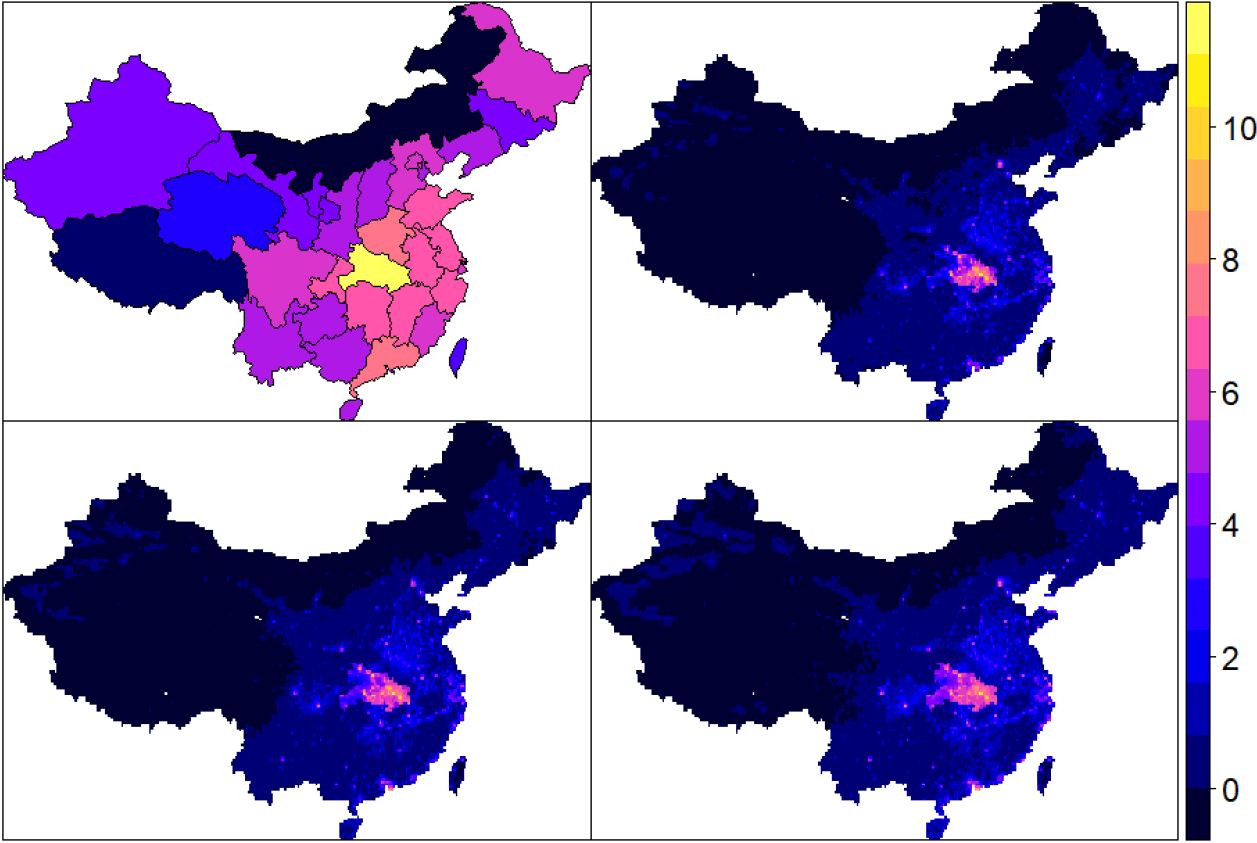
JHU original COVID-19 data and estimated cases (January-February 2020) with downscaling (natural logarithm scale) The maps show the original dataset of JHU of (natural log) COVID-19 infected cases for January and February 2020 at province-level in China (*top-left*), along with an estimation of the (natural log) infected cases based on the downscaling approach with lower bound (*top-right*), mean (*bottom-left*), and higher bound (*bottom-right*) predictions at 5 km grid-cell.

### 3.3. Comparing model predictions with disaggregated datasets at district-level

Since the reported cases from the disaggregated datasets are reported at city-level, they are likely to include cases that occur in the neighbourhood of the reported locations. To ease their comparison with the predicted cases from the downscaling approach, we compare the data at district level, which include 340 districts corresponding to one administrative level below province in China (Taiwan districts are merged into one single entity).

Fig 6 shows the number of (natural log) COVID-19 infected cases for January and February 2020 at district-level (340 districts) in China from the Paper (*left*), Xu et al. (*center*), along with an estimation of the (natural log) mean infected cases based on the downscaling approach (*right*). Figs 7 and 8 show the number of (natural log) COVID-19 infected cases for January and February 2020 at district-level (340 districts) in China from the Paper (*blue triangles*), Xu et al. (*red points*), along with an estimation of the (natural log) mean (*white squares*), 95% credible intervals (*grey segments*) of the infected cases based on the downscaling approach. For each district, the color of the symbol is faded for the corresponding dataset (Xu et al. or The Paper) that exhibits the most distant values (in absolute terms) to the predicted (natural log) mean cases from the downscaling approach.

**Figure 6:**
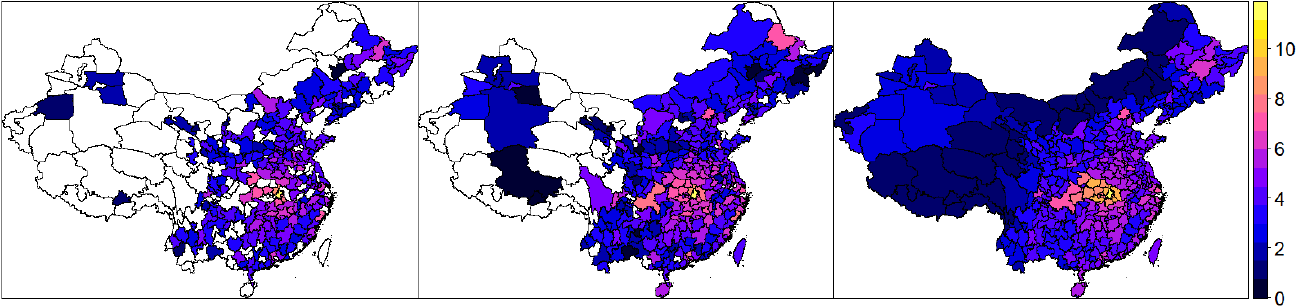
The Paper, Xu et al., and estimated cases (January-February 2020) with downscaling (log scale) in Chinese districts. The maps show the number of (log) COVID-19 infected cases for January and February 2020 at district-level (340 districts) in China from the Paper (*left*), Xu et al. (*center*), with blank areas corresponding to locations without observations provided by the corresponding disaggregated dataset. *Right*: estimated (natural log) mean infected cases based on the downscaling approach.

**Figure 7:**
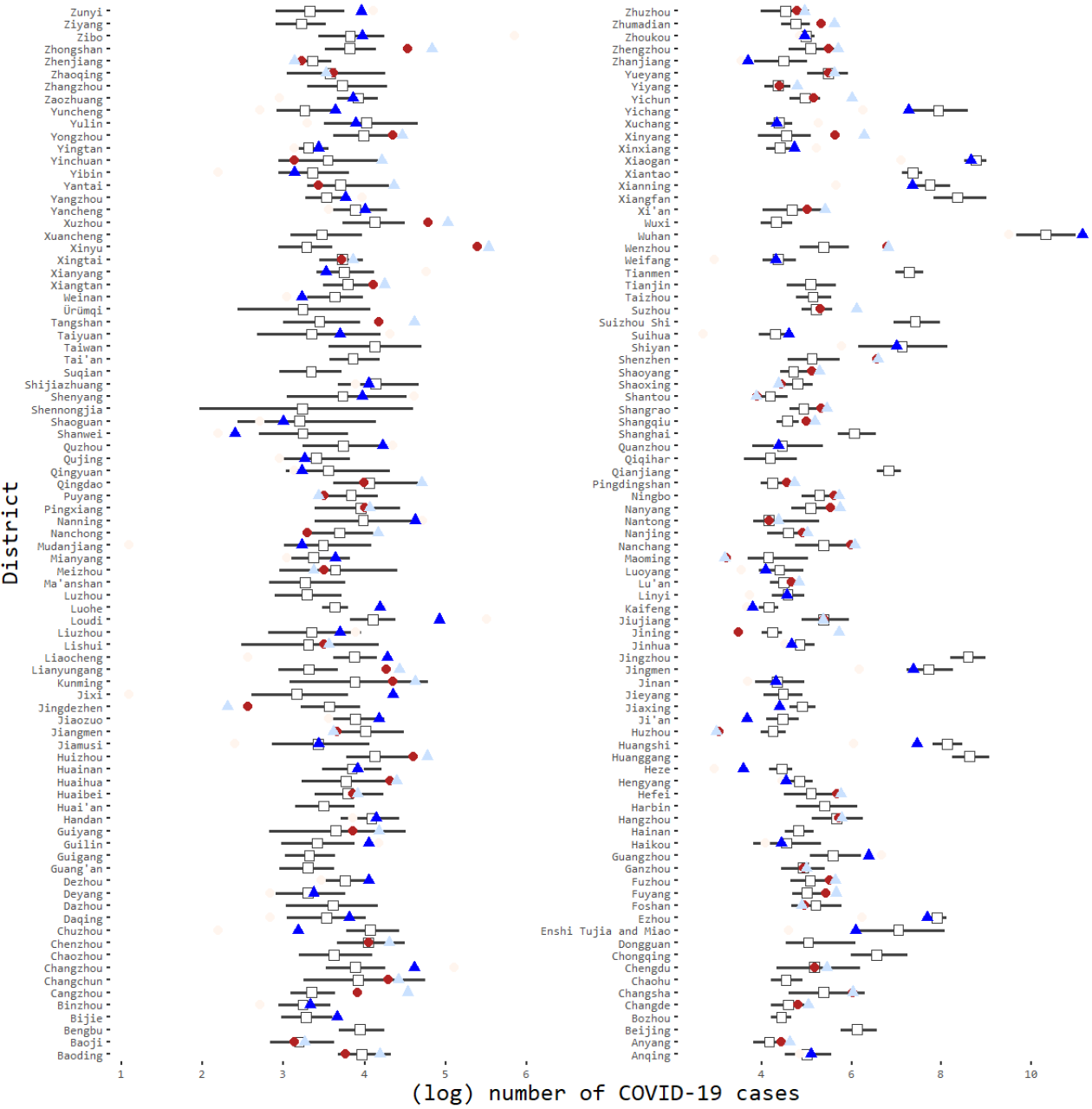
Assessment of consistency of reported cases from The Paper and Xu et al. with estimated cases from downscaling (natural log scale) JHU data in high-impacted districts. The figure shows the number of (natural log) COVID-19 infected cases for January and February 2020 at district-level (340 districts) in China from the Paper (*blue triangles*), Xu et al. (*red points*), along with an estimation of the (natural log) mean (*white squares*), 95% credible intervals (*grey segments*) of the infected cases based on the downscaling approach. For each district, the color of the symbol is faded for the corresponding dataset (Xu et al. or The Paper) that exhibit values less close to the predicted (natural log) mean.

**Figure 8:**
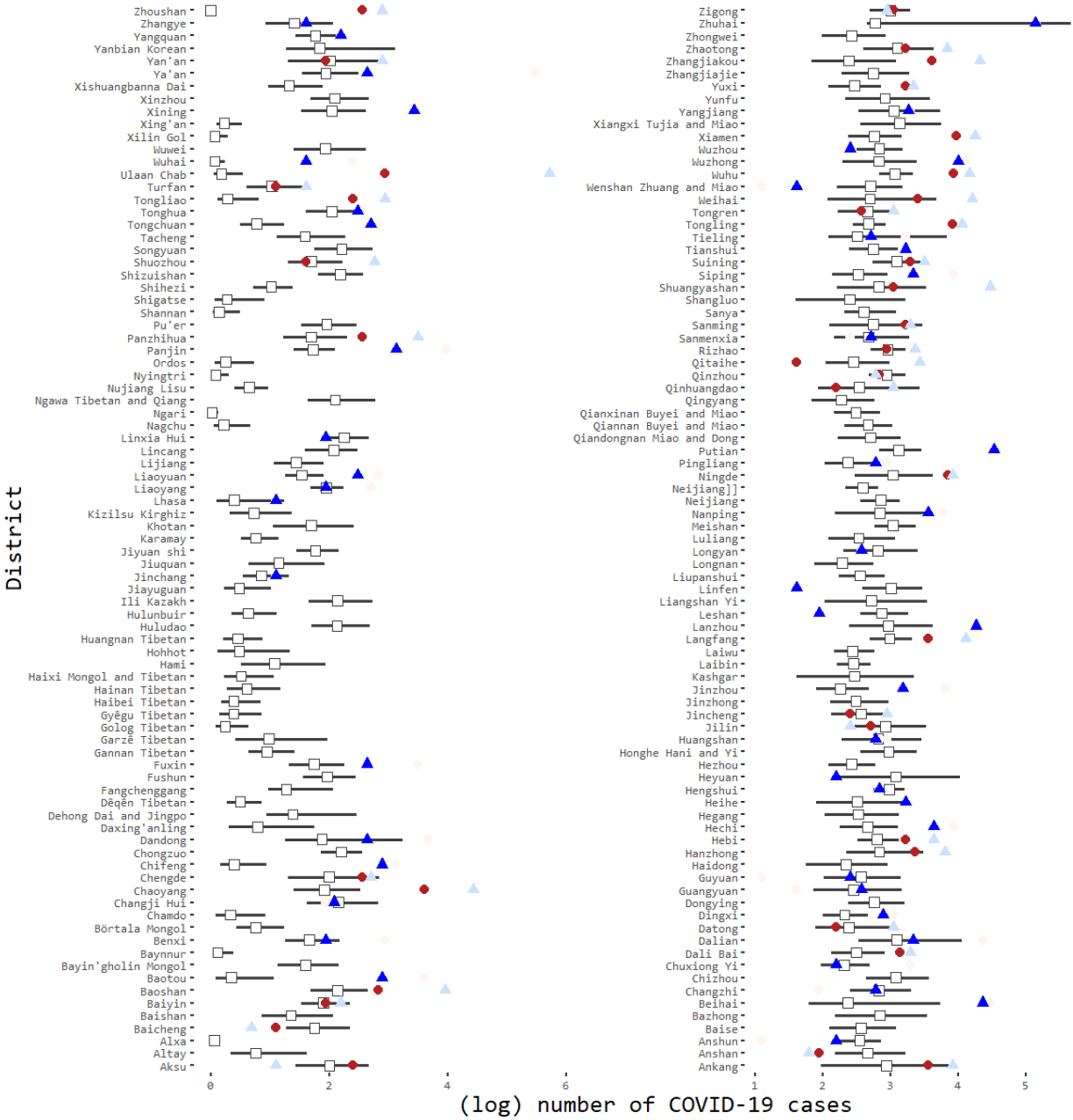
Assessment of consistency of reported cases from The Paper and Xu et al. with estimated cases from downscaling (natural log scale) JHU data in low-impacted districts. The figure shows the number of (natural log) COVID-19 infected cases for January and February 2020 at district-level (340 districts) in China from the Paper (*blue triangles*), Xu et al. (*red points*), along with an estimation of the (natural log) mean (*white squares*), 95% credible intervals (*grey segments*) of the infected cases based on the downscaling approach. For each district, the color of the symbol is faded for the corresponding dataset (Xu et al. or The Paper) that exhibit values less close to the predicted (natural log) mean.

Figs 7 and 8 show that in most districts, the values of the (natural log) reported cases from Xu et al. and the Paper lie within the 95% credible intervals of the predicted (natural log) cases based on the downscaling approach. This suggest that there is relative consistency between the investigated spatially disaggregated datasets (when data is provided) and the predictions from the downscaling model at district-level. We observe that the counts from Xu et al. tend to be closer to the predictive values from the downscaling approach. To further analyze potential discrepancies, we compute the absolute error (MAE) and the root mean squared error (RMSE) between the observed counts and the predictions based on several downscaling model specifications at district-level.

Table 1 shows the MAE and RMSE computed at district-level between each disaggregated dataset (Xu et al. and the Paper) and mean predicted values from downscaling models with 8 specifications (*Model spec*). The models differ by the use of: anthropogenic covariates (*Socio. cov*.), environmental covariates (*Env. cov*.), and i.i.d random polygon-specific effects (*i*.*i*.*d random*), and parameter tuning on the spatial parameters. We used default penalized complexity (PC) priors the spatial parameters with default thresholds *ρ min* and *σ max* and used alternative thresholds in model specifications (5-8).

**Table 1:** Performance metrics computed at district-level with several model specifications. Performance metrics computed to compare the discrepancies on the COVID-19 counts of the spatially disaggregated datasets from Xu et al. and The Paper with predicted (mean) values from the downscaling approach at district-level (340 districts) using different model specifications.

| Model spec. | Socio. cov. | Env. cov. | iid. random | $\rho$ min. | $\sigma$ max. | Nobs Xu | Nobs Paper | MAE Xu | MAE Paper | RMSE Xu | RMSE Paper |
| --- | --- | --- | --- | --- | --- | --- | --- | --- | --- | --- | --- |
| 1 | yes | yes | yes | 1 | 1.0 | 301 | 234 | 212.7 | 218.1 | 1283.7 | 2294.5 |
| 2 | yes | no | yes | 1 | 1.0 | 301 | 234 | 209.8 | 216.4 | 1266.4 | 2323.4 |
| 3 | no | yes | yes | 1 | 1.0 | 301 | 234 | 199.2 | 355.8 | 794.2 | 3738.1 |
| 4 | yes | yes | no | 1 | 1.0 | 301 | 234 | 305.2 | 162.8 | 4277.4 | 1284.3 |
| 5 | yes | yes | yes | 10 | 1.0 | 301 | 234 | 212.5 | 218.2 | 1285.0 | 2292.7 |
| 6 | yes | yes | yes | 5 | 1.0 | 301 | 234 | 212.4 | 217.3 | 1288.8 | 2287.4 |
| 7 | yes | yes | yes | 1 | 0.5 | 301 | 234 | 208.8 | 232.3 | 1165.5 | 2459.2 |
| 8 | yes | yes | yes | 1 | 5.0 | 301 | 234 | 211.6 | 222.2 | 1250.8 | 2339.4 |

Note that the data coverage varies among the disaggregated datasets. Xu et al. provides data in more districts (301) compared to The Paper (234). Missing data can be visualized in Fig 6 as blank areas. Data coverage is usually better in the East of China. The results of the analysis at district-level show that Xu et al. data have a larger data coverage and exhibit a number of reported cases closer to the model predictions compared to The Paper data. The predictive performance metrics MAE and RMSE are systematically lower with Xu et al. data (lower in 7/8 model specifications).

## 4. Discussion

The Bayesian downscaling model suggested in this paper made possible an assessment of discrepancies in the COVID-19 counts between databases aggregated at different spatial scales. By investigating reported cases from January to February 2020, the suggested approach allowed us to quantify differences in COVID-19 counts between two near real-time spatially disaggregated datasets and a reference province-level dataset at district-level in China. This data comparison would not be feasible with traditional statistical models. Furthermore, our analysis benefited from the Bayesian structure of the model, which provided a coherent framework to estimate and visualize uncertainty in the predictions across a fine spatial grid that covers our study area.

The method proposed in this paper exhibits several shortcomings. Our initial exploratory analysis remains descriptive and do not capture the processes behind the generated data. As such, it has only allowed us to identify potential data mismatches. However, the exploratory phase led us to investigate the observed data discrepancies in further detail. The second step of our analysis consisted of a comparison between reported coronavirus-infected cases and their locations gathered from two spatially disaggregated datasets (Xu et al. and The Paper^[16,45]^) and predictions from a downscaling approach using JHU data as reference province-level database^[15]^. Without reference data at a finer scale (city-level), we were not able to rigorously validate the results of the grid-cell predictions, and hence, identify data discrepancies with high confidence. Furthermore, our model did not distinguish age and gender classes of the population at risk. A potential refinement of the model informed by population structure could be used to better estimate the population at risk.

Assuming a well-specified model, the downscaling model is likely to have provided an accurate representation of the uncertainty^[23]^, which has allowed us to quantify deviations of the counts from the disaggregated databases with regard to the estimated mean and 95% credible intervals of the predicted counts from the downscaling model. To prevent the risk of overfitting, we built a relatively parsimonious model by selecting a few theoretically-relevant covariates and constraining them to be linearly associated with COVID-19 counts to keep the model as simple as possible.

Since in-sample predictions may not provide accurate predictive metrics in the presence of overfitting^[46]^, we selected the model based on the results of an out-of-sample iterative procedure that made predictions at grid-cell within each hold-out province from different model specifications. This procedure ensures that the resulting predictive metrics are based exclusively on data that has not been used in the fitting procedure. The model with the best predictive performance includes exclusively anthropegeic covariates, which brings evidence of the dominant role of human factors in the spread of COVID-19 ^[33]^.

A recent study that investigates the predictive performance of downscaling models exploring different contexts (various point data, aggregated areas sizes, and types of model misspecifications) suggests that predictive performance is likely to improve with a high number of data points and small polygon areas. If these conditions are not satisfied, predictions should remain accurate enough if the model is well-specified^[23]^. With a total of 33 polygons used to fit a relatively parsimonious model, we are confident that the predicted mean and uncertainty (95% credible intervals) should be accurate enough to provide reliable estimates of COVID-19 counts at district-level.

Despite the measures implemented to mitigate the effects of several major validity threats, our results remain dependent on the accuracy and reliability of the JHU reference data at province-level. Even at national and province-level, estimating the true number of positive cases of COVID-19 remains challenging. The number of individuals with a positive test result for the virus are inevitably smaller than the actual number of cases since a large proportion of infected people will not experience symptoms and may not get tested for the virus. Furthermore, the quality and efficiency of tests are subject to errors, which may lead to important discrepancies between reported and true counts^[34]^.

## 5. Conclusion

The analysis of spatially disaggregated COVID-19 data in January and February 2020 in China highlights important discrepancies in the counts of COVID-19 cases at a fine spatial scale. The results of an initial exploratory analysis led us to compare the observed differences in COVID-19 cases in China gathered from two spatially disaggregated datasets, Xu et al.^[16]^ and The Paper^[45]^, with fine-scale predictions using a Bayesian downscaling approach applied to a province-level data provided by Johns Hopkins University Center for Systems Science and Engineering (JHU)^[15]^. At district-level, COVID-19 counts from the spatially disaggregated datasets tend to be consistent with the range of plausible values of the predictions obtained by the downscaling model. We showed that, in comparison with The Paper^[45]^, Xu et al. data^[16]^ has a larger data coverage and the number of reported COVID-19 cases tend to exhibit a higher degree of consistency with the expected values of the predictions obtained from the downscaling model

The discrepancies in the counts of COVID-19 observed at province and district levels in China could also occur in other spatial and temporal frameworks. We believe that the data discrepancies highlighted in this case study are substantial enough to be considered by the providers of disaggregated data and the research community. As the COVID-19 pandemic continues to threaten various regions around the world, accuracy and reliability of the disaggregated COVID-19 data are crucial for governments and local communities to ensure rigorous assessment of the extent and magnitude of the threat posed by the virus and draw relevant conclusions. We hope that our analysis can help the data providers to identify at fine spatial scales potential data collection issues and remedy to them.

Governmental measures to mitigate the spread of the virus, such as quarantine, social distancing, and community containment are costly^[47]^. The efficiency of measures can be improved through evidence-based analysis grounded on reliable and accurate data. Given the observed data discrepancies at various spatial scales, we recommend that the providers of disaggregated COVID-19 data join efforts with the World Health Organization, as well as national and regional governments to build and maintain a comprehensive and reliable disaggregated database on COVID-19, which would help the research community and policy makers in their combat against the global threat posed by the virus.

## Data Availability

All data referred to in the manuscript is publicly available. The data sources are indicated in the method section.

## 6. Acknowledgment

We declare no competing interests. This work was supported by Zhejiang University Educational Funding (2020XGZX054), the National Natural Science Foundation of China under Grants (61825205 and 41601001), and The Royal Society, United Kingdom (NF171120).

## Supporting Information (SI)

This Section provides details on the data extraction and cleaning processes that has been performed to compare and select data on coronavirus. In addition, it provides further detail on the cross-validation procedure used to compare the predictive performance of disaggregated models. All operations (both computing and graphics) are carried out in the statistical software R^[48]^ and can be downloaded from this platform (link to be provided).

### 1. COVID-19 data selection

Data on coronavirus-infected cases are provided by various sources, including individual-level data from national, provincial, and municipal health reports, as well as additional information from online reports. A reference source of province-level data is provided by John Hopkins University CSSE (JHU)^[15]^ through an interactive web platform, whose data can be downloaded using github^[7]^. For cases in China, the sources used include Twitter feeds, online news services. The reported cases are confirmed with regional and local health departments, including centers for disease control and prevention of China, Taiwan, and Europe, the Hong Kong Department of Health, the Macau Government, and the World Health Organization (WHO), as well as city-level and state-level health authorities.

For the disaggregated datasets, we extracted city-level observations from the GitHub platform of the news agency Pengpai, which we refer to as “The Paper”^[6,45]^ (09 April 2020). We added city names in English, spatial coordinates of the cities (centroid), and dates in English. Furthermore, we removed events without or with wrong spatial coordinates. The cleaned dataset counts observations from 11 January 2020 to end of February 2020.

In addition to The Paper dataset, we extracted data from healthmap.org’s GitHub (09 April 2020), which includes COVID-19 cases in China in Hubei province^[8]^ and outside Hubei province^[9]^). We refer to this second dataset as Xu et al. data. The entire dataset (events without dates are removed) contains observations from January 18 to end of February 2020. The authors have provided the geolocalization of the data using google map and a variable that indicates the level of spatial accuracy, which can be used to subset the data. Xu et al. data has been aggregated through official government sources, peer-reviewed papers, and online reports. Various procedures have been applied by the authors to increase accuracy and comprehensiveness of the data. Therefore, we did not clean the original dataset since the provider used various robust procedures to ensure that the data is accurate and comprehensible, which include checking records and potential duplicates with peer-reviewed research articles.

### 2. Model validation

#### Out-of-sample predictions

Good in-sample performance may be the results of overfitting and are therefore not necessarily informative about the true performance of models. In order to assess the external validity of our model and to obtain more realistic estimations of performance metrics, we performed an out-of-sample cross-validation procedure by taking into account the spatial nature of the data and the model specifications^[49–51]^. While methods to evaluate out-of-sample performance of downscaling approaches have been developed (e.g.^[20,52]^), their relevance has been essentially drawn from the results of studies based on simulated data, which may not necessarily apply to our case study. Also, since province-level is the finer spatial resolution of our reference data (JHU data^[15]^), a crossvalidation approach based on spatial blocks^[53]^ would not be feasible.

As a result, we fit the downscaling models using data in all provinces except one (hold-out province), predicted the expected cases in the hold-out province, and reported performance metrics (RMSE, MAE) based on the expected cases from JHU. Through an iterative process, the data from the hold-out province is not used during the fitting process to ensure that the predictive performance of the models is assessed exclusively on new data. We performed the out-of-sample procedure on various model speficications, similarly to the insample procedure (see Table 1) except that we did not consider models with i.i.d random polygon-specific effects since we opted for a procedure that remove data from province in an iterative way. In this case, province-specific effects cannot be used to inform the model.

The model specifications (*Model spec*) are the following: (1) include all covariates, (2) include only anthropogenic covariates (*Socio. cov*.), (3) include only environmental covariates (*Env. cov*.), (4-7) using alternative penalized complexity (PC) priors on the spatial parameters *ρ min* and *σ max*. In total, this cross-validation computes predictive performance metrics for a total of 33 provinces multiplied by 7, which corresponds to a total of 231 runs. The results are illustrated in Table 2.

**Table 2:**
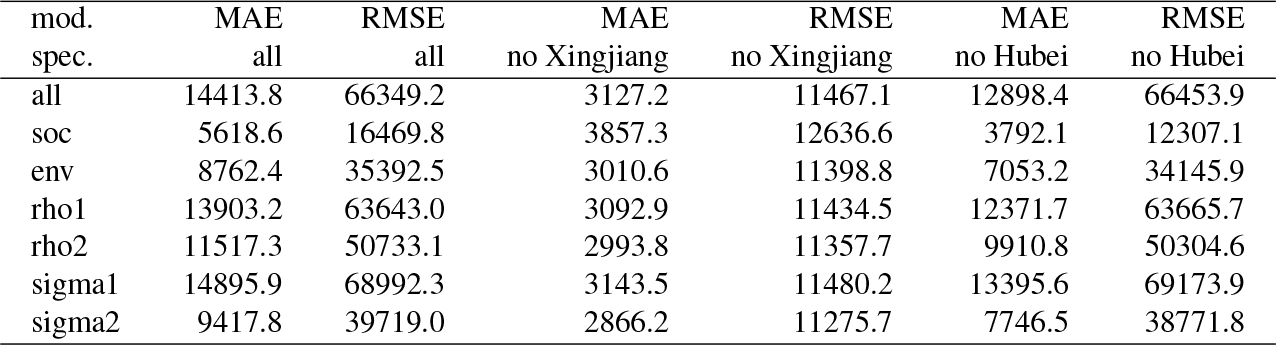
Performance metrics computed at province-level from an out-of-sample procedure. Performance metrics computed from an out-of-sample procedure (iteratively remove each of the 33 provinces) to compare different model specifications used to predict COVID-19 counts using a downscaling approach. We compare a total of seven model specifications (without i.i.d random effects): (1) include all covariates (*all*), (2) include only anthropogenic covariates (*soc*), (3) include only environmental covariates (*env*), and models that include all covariates but using alternative penalized complexity (PC) priors on the spatial parameters (*rho1,rho2,sigma1,sigma2*).

In Table 2, we report the MAE and RMSE (*MAE all, RMSE all*) for each model specification, based on the out-of-sample predictions in all provinces. Since the out-of-sample predictions in some provinces appear far more challenging in some provinces (see Xingiang and Hubei in Fig 9), we also present the MAE and RMSE results with predictions excluded in Xingjiang (*MAE no Xingjiang, RMSE no Xingjiang*) and Hubei (*MAE no Hubei, RMSE no Hubei*) to compare the model performance in different areas.

**Figure 9:**
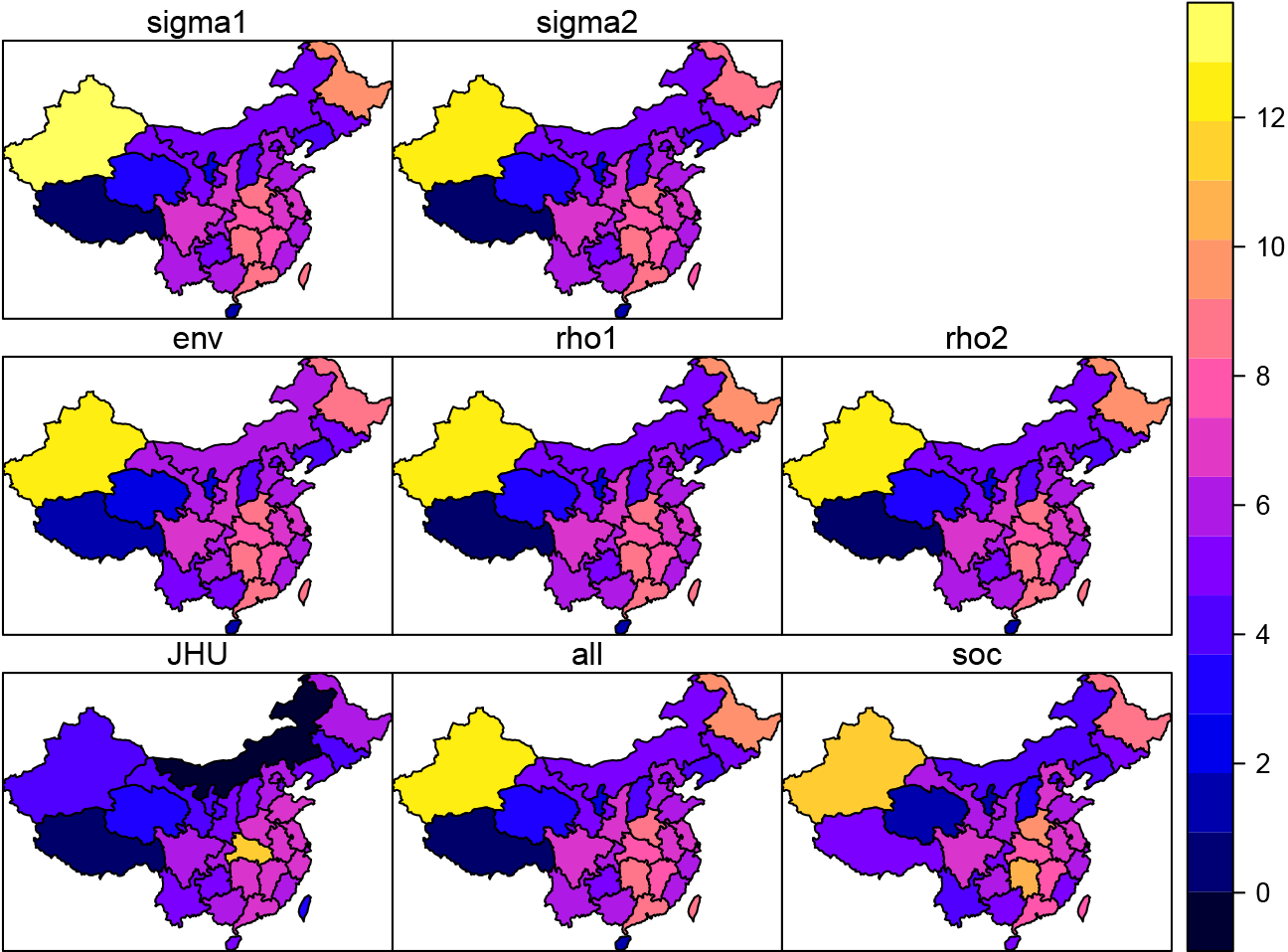
JHU original COVID-19 data and out-of-sample estimated cases (January-February 2020) with downscaling (natural logarithm scale). The maps show the original dataset of JHU of (natural log) COVID-19 infected cases for January and February 2020 at province-level in China (*bottom-right*), along with an estimation of the (natural log) infected cases based on the aggregation of all out-of-sample predictions in each of the 33 hold-out provinces from seven model specifications without i.i.d random effects: (1) include all covariates (*all*), (2) include only anthropogenic covariates (*soc*), (3) include only environmental covariates (*env*), and models that include all covariates but using alternative penalized complexity (PC) priors on the spatial parameters (*rho1,rho2,sigma1,sigma2*).

The results in Table 2 show that the model using only anthropogenic covariates (mod spec.: *soc*) has a better out-of-sample predictive ability in general, when we include predictions in all provinces (lowest *MAE all* and *RMSE all*) and also when we exclude predictions in Hubei province (lowest *MAE no Xingjiang* and *RMSE no Xingjiang*). If we exclude predictions in Xingjiang province, model *sigma2* with all covariates and alternative penalized complexity (PC) prior on the spatial parameter *sigma*_*max*_ = 5 shows the highest predictive performance. From these results, we select model *soc*, which has the best predictive performance across the different predictive contexts.

#### Insample predictions

Note that the out-of-sample predictions approach that remove in an iterative way each province (hold-out province) cannot be used to assess the predictive performance of models with province-specific i.i.d random effects. However, random effects can account for important differences we observe among provinces that cannot be informed by the covariates. Therefore, our final selected model is based on the selected specification (model using only anthropogenic covariates (mod spec.: *soc*)), by including an i.i.d random effects to account for province-specific characteristics.

To assess the insample predictive ability of the model, we compare the predicted incidence rate at province-level with the observed incidence rate computed as the number of reported COVID-19 cases from JHU^[15]^ divided by the population size^[30]^ of the province. For each province, the average incidence rate predicted by the downscaling is given by the sum of the mean predicted incidence rate multiplied by population size for each grid-cell within the province. A brief look at the results in Table 3 indicates that in most province, the predicted incidence rate (per 1,000) is equal to the observed incidence rate (per 1,000) when rounded at 3 digits. We can conclude that the in-sample predictions of the model are of sufficient accuracy.

**Table 3:** Results of insample predictions of the incidence rates (per 1,000) at province-level. Comparison of the observed incidence rate (per 1,000) at province-level (*second row*) with the average predicted incidence rate (per 1,000) (*third row*). The predictions are carried out in a total of 33 Chinese provinces are included.

| Province | 1 | 2 | 3 | 4 | 5 | 6 | 7 | 8 | 9 | 10 | 11 | 12 | 13 | 14 | 15 | 16 |
| --- | --- | --- | --- | --- | --- | --- | --- | --- | --- | --- | --- | --- | --- | --- | --- | --- |
| Obs.rate | 0.016 | 0.020 | 0.020 | 0.008 | 0.003 | 0.013 | 0.005 | 0.004 | 0.025 | 0.004 | 0.012 | 0.013 | 0.065 | 1.086 | 0.015 | 0.000 |
| Pred.rate | 0.016 | 0.020 | 0.020 | 0.008 | 0.003 | 0.013 | 0.005 | 0.004 | 0.025 | 0.004 | 0.012 | 0.013 | 0.065 | 1.086 | 0.015 | 0.000 |

Table 3: **Results of insample predictions of the incidence rates (per 1,000) at province-level.**
| Province | 17 | 18 | 19 | 20 | 21 | 22 | 23 | 24 | 25 | 26 | 27 | 28 | 29 | 30 | 31 | 32 | 33 |
| --- | --- | --- | --- | --- | --- | --- | --- | --- | --- | --- | --- | --- | --- | --- | --- | --- | --- |
| Obs.rate | 0.008 | 0.020 | 0.003 | 0.003 | 0.011 | 0.003 | 0.006 | 0.008 | 0.017 | 0.003 | 0.006 | 0.003 | 0.010 | 0.003 | 0.000 | 0.004 | 0.022 |
| Pred.rate | 0.008 | 0.020 | 0.003 | 0.003 | 0.011 | 0.003 | 0.006 | 0.008 | 0.017 | 0.003 | 0.006 | 0.003 | 0.010 | 0.003 | 0.000 | 0.004 | 0.022 |

